# Using the Social Ecological Model to explore policy and institutional aspects of the school environment in relation to healthy eating and physical activity: An audit of female-only high schools in the Kingdom of Saudi Arabia

**DOI:** 10.64898/2025.12.19.25342648

**Authors:** Sarah Aldukair, Jayne V. Woodside, Khalid Almutairi, Laura McGowan

## Abstract

**Objective:** In the Kingdom of Saudi Arabia (KSA), non-communicable diseases (NCDs) cause 73% of deaths, with high body mass index (BMI) as the leading risk factor. Among adolescents, 79% reported physical inactivity (PA), 30% and 10% were living with overweight/obesity and prediabetes, respectively. Little is known about the influence of school environments in shaping healthy eating (HE) and PA behaviours. This study addressed this gap through two steps: (1) Documentary review of obesity prevention policies, (2) Assessing the school environment in relation to HE and PA.

**Design:** The documentary review consisted of two parts; Part 1: Web-searching literature, hand-searching targeted websites, and stakeholder consultations to confirm the literature findings. Part 2: School-based environmental audit employing the ISCOLE tool and the Saudi Ministry of Education Regulations for School Canteens policy.

**Setting:** Three female high schools were selected from differing economic deprivation levels in Riyadh, KSA.

**Participants:** ISCOLE questionnaires were completed by the researcher in conjunction with a staff member; photo-documentation and observations were made.

**Results:** Seventeen obesity prevention policies were identified; stakeholder consultations identified one additional policy document, and assessment highlighted challenges in policy implementation. The audit revealed notable differences between schools in HE and PA provisions

**Conclusion:** In KSA, female high schools face significant challenges regarding a school environment that supports both HE and PA. These challenges include the absence of HE provisions and lack of suitable PA facilities, coupled with a gap between policy and practice. Understanding the school context will help support the development of future obesity prevention school-based interventions.

## 1. Introduction

In the Kingdom of Saudi Arabia (KSA), the burden of mortality and morbidity is mainly caused by non-communicable diseases (NCDs), which are estimated to account for 73% of all deaths (1). With high body mass index (BMI) being the leading risk factor for disease, and NCDs being the leading causes of disease burden among Saudi youths (2). This trend is of serious public health concern given that KSA has a youthful population, with 67% of the population aged less than 35 years, and approximately one quarter of the population (25%) are adolescents who fall in the age group of 15-19 years (3). Even though youth make up a significant percentage of KSA’s population, they have received minimal attention regarding their health status (4). With rapid societal developments occurring in KSA in relation to urbanization, a shift in dietary habits and food choices has emerged (5). Physical inactivity, skipping breakfast, low consumption of fruits and vegetables, high consumption of calorie-dense food and sugar-sweetened beverages are widespread among Saudi adolescents and especially among females (5–6), contributing to rising rates of obesity (7–9). Among Saudi adolescents aged 15–29 years, 79% reported insufficient PA, 30% were living with overweight or obesity, pre-diabetes and raised cholesterol affected 10% and 39%, respectively (10). KSA is a high-income country with abundant resources, and investing in its youthful population’s health is suggested to prevent future health burdens (4). Whilst obesity is recognized as a complex, multi-factorial chronic condition (11), the Saudi Guidelines for Obesity recommend school-based interventions (SBIs) for improving adolescent health (12), yet little is known about how the school environment influences risk factors for obesity, such as HE and PA, which play an important role in obesity prevention. Understanding the school setting through a school-based environmental audit can provide insight into the barriers and enablers of obesity prevention strategies in schools (13), which can inform the design of SBIs. One such tool is the International Study of Childhood Obesity, Lifestyle and the Environment (ISCOLE) questionnaire, which assesses the school environment in relation to HE and PA. The reliability and feasibility of this tool have been tested across 256 schools in 12 different countries (13), capturing both policy and environmental aspects aligned with the socio-ecological model (SEM) (14). Prior ISCOLE findings have revealed that variations in school environments shaped students’ HE and PA behaviours; across 12 countries, supportive school environments with school policies and access to PA facilities reported better obesity prevention outcomes (15). Moreover, more than 100 peer-reviewed papers have been published using the ISCOLE tool (15). Therefore, the ISCOLE tool can be used to conduct reliable environmental audits across international school settings. While other audit tools exist, such as the Centers for Disease Control and Prevention School Health Index and the World Health Organisation’s Health Promoting Schools framework, they lack international validation (16–17). Previous studies in Saudi schools have revealed the high availability of unhealthy food items. For instance, Aldubayan and Murimi examined male schools’ food offerings using the Institute of Medicine standards, and found energy-dense, pre-packaged foods, with almost all schools offering cakes and confectionaries (18). Moreover, Togoo et al. found high availability of cariogenic and high-calorie items in primary schools (19). Futhermore, Alsiwat et al. examined female primary schools using the MOE Regulations for School Canteens and found low policy compliance (20). While these studies offer some insights into school food environments in KSA, compliance remains underexplored, especially in female school settings. This study addressed this gap through examining the intersection between existing policy and practice, by conducting a documentary review of obesity prevention policies and an in-depth school-based environmental audit in three female-only high schools in Riyadh from different levels of economic deprivation, to examine the extent to which the school environment supports HE and PA behaviours. This study employed the SEM as a framework to explore the wider environmental aspects of the school environment and how they may influence HE and PA behaviours, as well as exploring the policy context (14). Using the SEM enabled the exploration of both the broader policy context and the environmental factors in school settings, providing a comprehensive understanding of how school structures and policies interact to support or hinder HE and PA behaviours.

## 2. Methods

Methods are presented in two parts: i) documentary review of existing obesity prevention policies in KSA, and ii) a school-based environmental audit.

For part i), a systematic search strategy was devised which included web-based searches, targeted hand-searches of the KSA MOH and MOE websites, and consultations with institutional stakeholders (MOH and MOE representatives).

For part ii), a school-based environmental audit was conducted focusing on three key areas: HE provisions and PA facilities within school settings, and obesity prevention policies. This was conducted by using a modified version of the International Study of Childhood Obesity, Lifestyle and the Environment (ISCOLE) school environment questionnaire (13), coupled with a compliance checklist from MOE’s Regulations for Canteens policy (21).

### 2.1 Documentary review of existing obesity prevention policies and strategies in KSA (Part i)

A documentary review of existing obesity prevention policies in KSA serves as a comprehensive examination of the country’s current approach to tackling the obesity epidemic, with a focus on obesity prevention within the school setting. Relevant policy documents were identified through web-searching the literature and targeted website searches and verified through stakeholder consultations with representatives from the MOH and MOE. It consisted of the three following steps:

#### 2.1.1 Web-searching the literature

A systematic grey literature search was conducted to find relevant policies and strategies related to obesity prevention. This web-search was guided by the systematic public health policy review framework by Godin et al. (22), which involved searching the grey literature using custom “advanced Google search”. The keywords used to search Google Advance database are illustrated in Table 1.

**Table 1:**
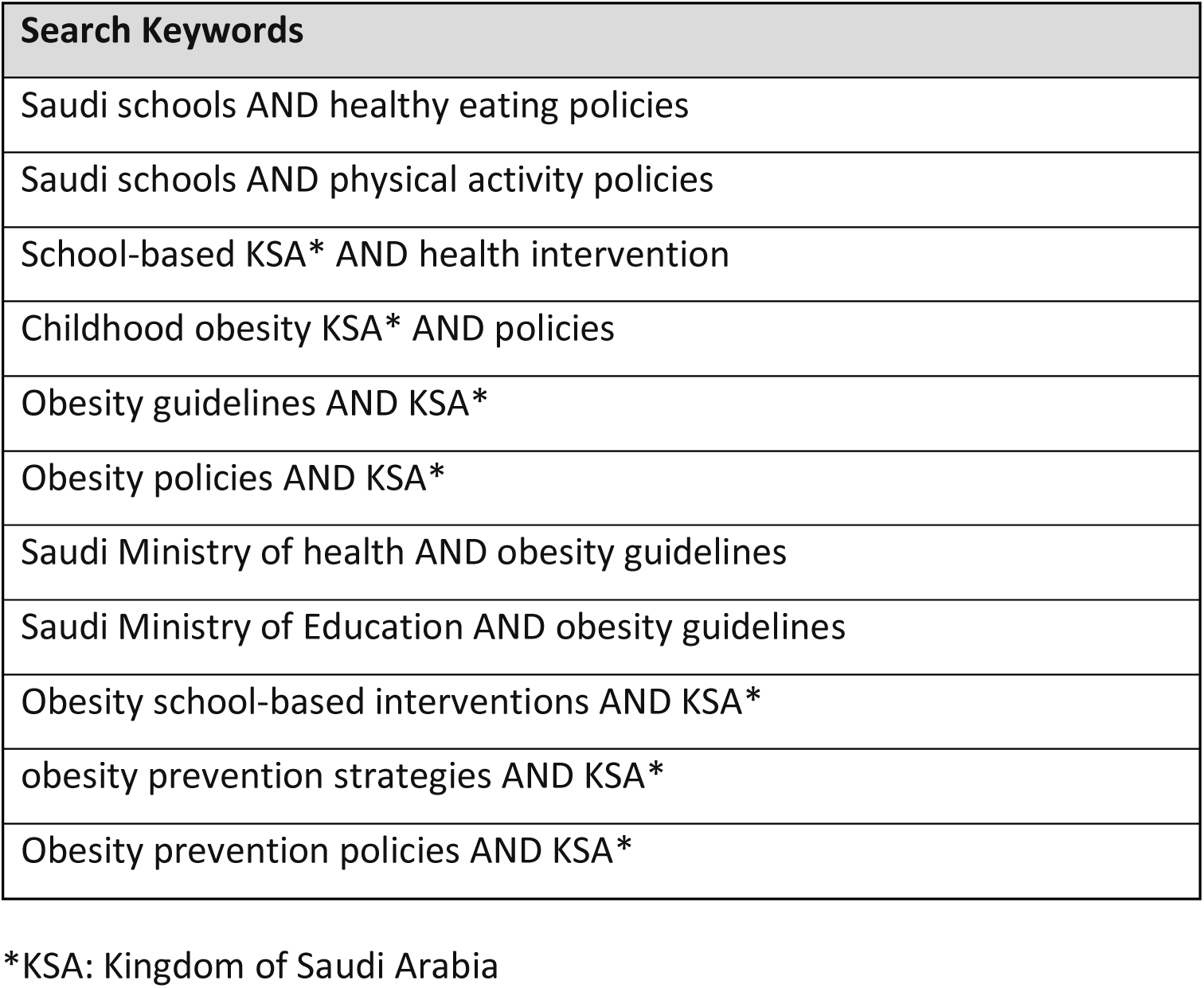
Search Keywords.

#### 2.1.2 Hand-searching targeted websites (MOH and MOE websites)

Keywords are summarized in Table 2 for MOH and MOE websites. For each keyword, the first 100 webpages were screened, consistent with the number screened in Google Advanced searches.

**Table 2:** Ministry of Health and Ministry of Education website keywords.

| Ministry of Health and Ministry of Education website keywords |
| --- |
| Obesity prevention |
| Adolescent obesity |
| Adolescent health |
| School-based health awareness program |
| Nutrition programs |
| School health |
| Obesity policies |

#### 2.1.3 Stakeholder consultations

Findings from web-searching the literature and hand-searching targeted websites were confirmed through placing a 20-25 minute phone consultations with MOH and MOE representatives to confirm relevant obesity prevention policies and strategies, particularly those targeting schools. Ethical approval was not required, as this process involved verification of publicly available information.

### 2.2 School-based environmental audit (Part ii)

A school-based environmental audit was conducted using the ISCOLE school environment questionnaire (13) (See Supplementary File S1), coupled with the Saudi MOE Regulations for School Canteens policy checklist (21) (See Supplementary Table S2), to examine the schools’ built environments in relation to HE and PA. Three female-only high schools in Riyadh, representing different levels of economic deprivation levels (high, medium, low) were purposively selected. Data collection included interviews with principals, direct observation, and photo documentation of food and PA facilities. The researcher sent e-mails to school principals, then followed up with phone calls to ask for permission to conduct the school-based environmental audit. School principals were given information sheets with a detailed description of the study. After getting the principals’ verbal approval, consent forms were filled out, and interviews were scheduled to conduct the study in the three schools. Principals were interviewed to respond to questions from the ISCOLE school environment questionnaire; they were asked the questions, and they responded in an interview-like scenario. The researcher made some direct observations by visiting the school canteen and PA facilities, and photo-documented food and PA facilities. The ISCOLE questionnaire was minimally modified to fit the cultural context of Saudi schools. Modifications are outlined in Table 3.

**Table 3:** ISCOLE questionnaire cultural adaptation.

| Question | Modification | Reason |
| --- | --- | --- |
| **C9** | -The following words were removed; varsity athletics from (C9.a) and intramural athletics including dance from (C9.b) | -Not applicable to schools in KSA* |
| **C11** | -The terms: “interschool or intramural athletics program” were removed from the question and were replaced by “school” instead.<br>-From the list of PA types offered in the school setting, skiing and ice hockey were removed, | -Not applicable to schools in KSA*<br>-Skiing and ice hockey were never practiced in KSA* |
| **C11-C15** | - “Grade 4” was replaced with “grades 10-12” | -To represent high school grades surveyed in the study |
| **C14** | -The terms “province/territory/state” were replaced with “the Ministry of Education” | -Schools in KSA* are under the jurisdiction of the Ministry of Education |
| **C17** | -This question was about promoting active transportation to school- it was entirely replaced with the following question: “What kind of transportation do students use to arrive to their | -Because Riyadh city is not pedestrian-friendly, and no one walks or cycles to school as a means of |
|  | school?" and the following answers: a. car, b. bus, c. other | transportation. The city inhabitants heavily rely on cars for transportation (19) |
| **D18** | - "Skating rink/arena" was removed from the checklist of physical activity facilities | -Skating is not a common activity in KSA*, especially in schools. |
| **E26** | - "During the past 12 months, have any of the following items been sold as part of fundraising for any school organisation?" was removed from the questionnaire | -A comprehensive checklist of recommended and prohibited food items was already included in the audit, in alignment with the Ministry of Education's Regulations for School Canteens. |
| **F27** | -The term "excessive drinking" was removed from the checklist of issues in the surrounding neighborhood of the school | Drinking alcohol is strictly prohibited in KSA* |

\*KSA: Kingdom of Saudi Arabia

## 3. Results

### 3.1 Documentary review of existing obesity prevention policies and strategies in KSA (Part i)

#### 3.1.1 Web-searching the literature

The number of results, the number of results screened, and number of relevant webpages are shown for each set of keywords in Table 4. Web-searching the literature resulted in identifying five national policies and strategies regarding obesity prevention in schools presented in Table 5.

**Table 4:** Keyword results from web-searching the literature.

| Search Keywords | Number of results | Number of results screened | Relevant webpages |
| --- | --- | --- | --- |
| Saudi schools AND healthy eating policies | 4,630,000 | 100 | 9 |
| Saudi schools AND physical activity policies | 57,100,000 |  | 12 |
| School-based KSA* AND health intervention | 11,900,000 |  | 19 |
| Childhood obesity KSA* AND policies | 3,500,000 |  | 21 |
| Obesity guidelines AND KSA* | 1,310,000 |  | 23 |
| obesity policies AND KSA* | 5,460,000 |  | 18 |
| Saudi Ministry of health AND obesity guidelines | 304,000 |  | 8 |
| Saudi Ministry of Education AND obesity guidelines | 207,000 |  | 6 |
| Obesity school-based interventions AND KSA* | 657,000 |  | 16 |
| obesity prevention strategies AND KSA* | 737,000 |  | 23 |
| Obesity prevention policies AND KSA* | 2,960,000 |  | 21 |

\*KSA: Kingdom of Saudi Arabia

**Table 5:** Documentary review results (web-searching)

| Documentary review results (web-searching) |  |  |
| --- | --- | --- |
| Policy/Strategy | Year | Details |
| **Ministry of Education - Regulations for School Canteens** | - Established in 2004<br>- Updated in 2013 then in 2019 | - Offering healthy food options<br>- Limiting unhealthy snacks<br>- Complying with food safety standards |
| **Ministry of Education - Assigning a health advisor to each school** | 2008 | - Promoting health education and healthy school environment<br>- Performing health screening for students |
| **Ministry of Education and Ministry of Health - RASHAKA initiative** | 2017 | - Reducing obesity rates by 5% |
| **Ministry of Health - Healthy Schools Program** | 2018 | - Improving the health of students according to the WHO school health initiative |
| **Ministry of Health - Saudi Guidelines for the Prevention and Management of Obesity** | 2016 | - Evidence-based recommendations for addressing obesity in both adults and children |

##### Regulations for School Canteens

The MOE established the Regulations for School Canteens in 2004, updated in 2013 and 2019, to promote HE habits by offering healthy food options and restricting unhealthy food items (21).

##### Assigning health advisors

In 2008, the MOE assigned a health advisor to each school to promote health education, ensure a healthy school environment, and perform health screenings.

##### The *RASHAKA* initiative

In 2017, the MOH and MOE initiated the *RASHAKA* program, a school-based initiative aiming to reduce obesity rates by 5% among school-aged children over five years. The program targets students, parents, and teachers to improve HE and PA behaviours, enhance school environments, provide healthier food options, and offer preventive and curative services, including obesity screening and referral to primary healthcare (23).

##### The Healthy Schools Program

In 2018, the MOH established the Healthy Schools Program, aligning with the WHO school health initiative (24).

##### The Saudi Guidelines for the Prevention and Management of Obesity

The Saudi Guidelines for the Prevention and Management of Obesity provide evidence-based recommendations emphasizing HE and PA and the need for multi-sectoral approaches, though they lack detailed strategies targeting the school setting (12).

#### 3.1.2 Hand-searching targeted websites

Table 6 illustrates the number of results and results screened from the MOH and MOE websites. Hand-searching MOH (25) and MOE (26) websites identified webpages on school-based programs and resources supporting HE and PA. Each keyword returned less than 100 results as illustrated in Table 7.

**Table 6:** Number of results identified through hand-searching of Ministry of Health and Ministry of Education websites.

| Hand-searching Ministry of Health website |  |
| --- | --- |
| Keyword | Number of results/results screened |
| Obesity prevention | 7 |
| Adolescent obesity | 1 |
| Adolescent health | 11 |
| School-Based Health Awareness Program | 2 |
| Nutrition programs | 2 |
| School health | 71 |
| Obesity policies | 1 |
| Hand-searching Ministry of Education website |  |
| Keyword | Number of results/results screened |
| Obesity prevention | 6 |
| Adolescent obesity | 2 |
| Adolescent health | 23 |
| School-Based Health Awareness Program | 13 |
| Nutrition programs | 3 |
| School health | 18 |
| Obesity policies | 2 |

**Table 7:** Relevant webpages of hand-searching targeted websites.

| Website | Relevant webpage title |
| --- | --- |
| www.moh.gov.sa | School-Based Health Awareness Program |
|  | Calorie manual for weight reduction |
|  | Breakfast is essential for students |
|  | Training Courses to Activate School Health Programs and Initiatives |
|  | «Healthy Schools» Program Launched to Improve Student Health |
|  | Contest for Schools in “*Wa3i*” Award Second Season |
| www.moe.gov.sa | School health |

The school-based health awareness program, a weight-loss calorie guidebook, breakfast awareness campaigns, school health initiative training courses, the Healthy Schools Program, and the *“Wa3i”* school health competition were among the MOH webpages. Information on School Health, aiming to promote healthy behaviours within the school community and provide a safe learning environment were included among the MOE webpages.

#### 3.1.3 Stakeholder consultations

A representative from KSA MOH, within MOH’s policy development team, was interviewed to verify the policies and documents found from web-searching the literature and hand-searching targeted websites as described above. This consultation confirmed the policies found and yielded one additional policy document: “Overweight and obesity in KSA: consequences and solutions” (27). The document also included school-based policies that confirm previous web-searching findings; MOE’s Regulations of Health Conditions for School Canteens (21), and the RASHAKA collaborative initiative between MOE and MOH (23). One chapter of the document describes obesity prevention strategies and policies in KSA, including fiscal policies, nutrition labeling, food and beverage reformulations, marketing restrictions of unhealthy food and beverages, and eight obesity-related national strategies summarized in Table 8.

**Table 8:** National obesity-related strategies relevant to school settings.

| National Strategy | Relevance to the school setting |
| --- | --- |
| 1. Vision 2030 (Quality of Life Program) | -Promotes community-wide wellness and physical activity, with a focus on improving health infrastructure, which can support schools in implementing physical activity programs.<br>-Encourages cross-sector collaboration, for example between the Ministry of Education and Ministry of Health, which supports school-based health promotion. |
| 2. Obesity Control and Prevention Strategy (2020–2030) | -Supports early intervention through schools, including improving school meals, physical education, and nutrition literacy.<br>-Integrates obesity prevention content into school curricula and policies. |
| 3. Obesity Control Program Strategy (2014–2025) | -Encourages stakeholder engagement in schools |
|  | -Recommends school-based screening and monitoring of body mass index for early obesity prevention |
| **4. Healthy Food Strategy (2018–2030)** | -It targets school canteens and improves food quality in school meals |
| **5. Diet and Physical Activity Strategy (2014–2025)** | -Supports active school environments |
| **6. National Strategy for Prevention of Non-Communicable Diseases (2014–2025)** | -Considers schools as important environments for Non-Communicable Diseases risk reduction |
| **7. National Executive Plan for Diabetes Control (2014–2025)** | -Considers schools as important environments for increasing awareness about diabetes risk factors (low physical activity, poor diet) |
| **8. National Strategy for Prevention of Cardiovascular Disease (2010–2020)** | -It emphasized early lifestyle interventions in schools to reduce long-term risk of cardiovascular diseases<br>-It promoted healthy eating habits |

Consultations with an MOE representative confirmed the existence of the Regulations for School Canteens policy and the *RASHAKA* initiative but noted that implementation is often neglected due to limited monitoring and follow-up.

A summary of strategies, policies, and programs initiated by governmental entities to tackle the obesity epidemic in KSA as a result of the documentary review can be found in Table 9.

**Table 9:** Governmental obesity prevention strategies, policies, and programs in KSA.

| Strategy/policy/program | Implementation Date | Stakeholder | Target group | Evaluation | Found via |
| --- | --- | --- | --- | --- | --- |
| **Regulations for School Canteens** | Established 2004, updated in 2013 and 2019 | Ministry of Education | School-aged children | Not reported; one compliance study exists (20) | 1. Web-searching the literature<br>2. Stakeholder consultations (with Ministry of Education/Ministry of Health representatives) |
| **Assigning a health advisor to each school for health promotion** | 2008 |  | School-aged children | Not reported | Web-searching the literature |
| **Obesity control program (Saudi guidelines for the Prevention and Management of Obesity)** | 2016 | Ministry of Health | Guidelines for physicians to prevent and manage obesity among the | Not reported | Web-searching the literature |
|  |  |  | Saudi population |  |  |
| **RASHAKA initiative** | 2017, over a 5-year period | Ministry of Education/Ministry of Health | school-aged children, their parents and teachers | Not reported | 1. Web-searching the literature<br>2. Stakeholder consultations (with Ministry of Education/Ministry of Health representatives) |
| **Healthy Schools Program** | 2018 | Ministry of Health | School-aged children | Not reported | Hand-searching targeted websites (Ministry of Health) |
| **Fiscal policies** | 2017 and 2019 | General Authority of Zakart, Tax, and Customs | Saudi population | Evaluated partially:<br>-Reduction in national sales<br>-No health impact evaluation studies | Stakeholder consultation (Ministry of Health) |
| **Nutrition-labeling policies** | 2018-2020 | Saudi Food and Drug Authority | Saudi population | Not reported |  |
| **Food and beverage product reformulations** | 2018-2020 |  | Saudi population | Not reported |  |
| **Marketing restrictions of unhealthy foods and beverages** | 2022 |  | Saudi population | Not reported |  |
| **Vision 2030 (quality of life program)** | 2016-2030 | Vision realization office | Saudi population | Not reported |  |
| **Obesity control and prevention strategy** | 2020-2030 | Public health authority | Saudi population | Not reported |  |
| **Obesity control program strategy** | 2014-2025 | Ministry of Health | Saudi population | Not reported |  |
| **Healthy food strategy** | 2018-2030 | Saudi Food and Drug Authority | Saudi population | Not reported |  |
| **Diet and physical activity strategy** | 2014-2025 | Ministry of Health | Saudi population | Not reported |  |
| **National strategy for prevention of NCDs** | 2014-2025 | Ministry of Health | Saudi population | Not reported |  |
| **National executive plan for diabetes control** | 2014-2025 | Ministry of Health | Saudi population | Not reported |  |
| **National strategy for prevention of CVDs** | 2010-2020 | Ministry of Health | Saudi population | Not reported |  |

### 3.2 The school-based environmental audit using the modified ISCOLE tool (Part ii)

As outlined in Table 10, the school-based environmental audit findings varied across the three schools when assessed using the modified ISCOLE questionnaire. In the policies section, the high deprivation school did not have any written policies or practices regarding HE or PA; the middle and low deprivation schools both reported policies that were ‘under development’, however none were finalized or implemented.

**Table 10:** School-based environmental audit results.

|  | High deprivation school | Middle deprivation school | Low deprivation school |
| --- | --- | --- | --- |
| **Written policies and practices relating to healthy eating and physical activity** | No policies or practices | Under development:<br>○ Making the physical education curriculum practical<br>○ Using the free time for PA<br>○ Healthy eating programs under the supervision of a health advisor | Under development (cannot disclose until complete) |
| **Presence of a committee that offers guidance on the development of policies and practices relating to healthy eating and physical activity** | Not present | Present | Present |
| **Types of physical activity offered in school** | No physical activity offered in school |  |  |
|  | **High deprivation school** | **Middle deprivation school** | **Low deprivation school** |
| **Time mandated by the Ministry of Education for physical activity** | No specific amount mandated | 30 minutes/week | 30 minutes/week |
| **Transportation used by students to reach school** | Car |  |  |
| **Type of physical activity facilities in offered** | No facilities available | Large room suitable for PA | ○ Outdoor paved area for physical activity<br>○ Soccer field |
| **Facility to buy food and drinks** | No canteen or shop available | School shop | School shop |
| **Offering healthy food choices at a subsidized price** |  | No | No |
| **Offering healthy eating promotional materials** | No | No | Yes (posters) |
| **Healthy eating activities/programs** | No | Healthy breakfast day (once or twice a year) | Healthy breakfast day (once or twice a year) |
| **Health promotion for healthy eating** | No | Media literacy on topics relating to healthy eating | Media literacy on topics relating to HE |
|  | **High deprivation school** | **Middle deprivation school** | **Low deprivation school** |
| **Unallowed food items present in school canteen as per the Ministry of Education policy** | No canteen or shop available | ○ Nectar juices<br>○ Popcorn<br>○ Chocolates<br>○ Croissants | ○ Nectar juices<br>○ Popcorn<br>○ Chocolates<br>○ Croissants<br>○ Potato chips |
| **Recommended food items present in school canteen as per the Ministry of Education's policy** |  | ○ Bottled water<br>○ Sandwiches with healthy fillings<br>○ Packaged date biscuits | ○ Bottled water<br>○ Sandwiches with healthy fillings<br>○ Milk/life-long milk |
|  | **High deprivation school** | **Middle deprivation school** | **Low deprivation school** |
| **Food presentation at school canteen** | No canteen or shop available | ○ No promotions or discounts offered<br>○ Food items are presented to students in school shop shelves and trays on a big table<br>○ Croissants and sandwiches are pre-packaged<br>○ Chocolates are the most prominent item sold<br>○ Students buy food from the school shop window | ○ No promotions or discounts offered<br>○ Food items are presented to students on school shop table<br>○ Croissants and sandwiches are pre-packaged<br>○ Students buy food from the school shop window |
| **Prices in comparison with local shops** |  | More expensive | Same price |
| **Proportion of students who rely on school canteen** | 0% | 50% | 60% |
| Proportion of students who rely on food from home (estimates from principal) | 100% | 50% | 40% |

Similarly, a committee responsible for guiding the development of policies and practices related to HE and PA was reported to be present in both the middle and low-deprivation schools but was absent in the high-deprivation school.

In the low deprivation school, a small area dedicated to soccer was identified (See Supplementary Figure S3). However, the vice principal commented that although it is present, no one uses it. Regarding the specific amount of PA mandated by the MOE, the high deprivation school did not have any time dedicated for PA, whereas the middle and low deprivation school had 30 minutes of PA per week. In all three schools, 100% of students used cars to reach their schools. In the high deprivation school, the principal reported the absence of PA facilities, and the researcher confirmed that by examining the premises. The school is small, with relatively less students than the usual number, and it is comprised of classrooms and a teacher’s room only. However, in the middle deprivation school, there is a large room suitable for PA, and in the low deprivation school, a small soccer field and an outdoor paved area were present. There are no food and drink facilities in the high deprivation school, not even a school shop. The principal commented that the students cannot afford to buy from the school shop, so they shut it down due to diminished revenues. However, in low and middle deprivation schools, a school shop is present. However, none of the schools offer healthy options at a subsidized price. Regarding offering HE promotional materials, the high and middle deprivation schools did not offer any, but the low deprivation school had promotional posters containing the food pyramid in Arabic (See Supplementary Figure S4). Both schools also offered healthy breakfast days once or twice a year, and HE awareness, unlike the high deprivation school which did not offer any.

The MOE regulations for school canteens checklist were applied on the middle and low deprivation schools only, since the high deprivation school did not have a school shop available. The unallowed food items present in both school shops were almost identical; nectar juices, pre-packaged croissants, chocolates, and popcorn. The only distinct unallowed item was crisps in the low deprivation school shop. Regarding the recommended food items, both schools offered sandwiches and bottled water. Milk was only present in the low deprivation school shop, and date biscuits were only present in the middle deprivation school shop. Both school shops had a window to buy from, and the food was mostly pre-packaged with no discounts or promotions.

When asked about pricing, the low deprivation school had the same prices as the local grocery store. However, the middle deprivation school had more expensive prices than the local grocery store. Both school shops were run by private companies, and their profit system is by percentage on each item sold. Photos of the school premises can be found in Supplementary Figures S4-S7.

## 4. Discussion

This study offers valuable insights into understanding how the school environment influences HE and PA behaviours in female-only high schools in Riyadh, KSA. To our knowledge, this is the first school-based audit study conducted in female-only high schools in Riyadh using the ISCOLE tool. It examines the policy and institutional aspects of the SEM in this context. The findings reveal notable discrepancies in implementing obesity prevention and health-related policies and practices, especially when comparing schools across different economic deprivation levels, highlighting the difficulties in aligning national obesity prevention strategies with the school environment. The documentary review revealed a gap between KSA’s national policies, and the obesity prevention practices within schools. Although the government has legislated a variety of obesity prevention policies, such as the MOE Regulations for School Canteens policy and the MOH campaigns, policy implementation at the school level remains weak and inconsistently enforced.

Having school nutrition policies that promote the availability and accessibility of healthy food are essential. These policies can create healthy school environments by providing healthy food options to students (28), yet in this school environmental audit it was found that canteens are not compliant with the MOE Regulations for School Canteens policy (21). The absence of compliance has weakened the effectiveness of this policy. This gap between policy and practice was consistently observed across schools of different deprivation levels, where unhealthy food items were present in school premises. For instance, the MOE Regulations for Canteens policy aims to improve the food sold in school canteens, by banning specific unhealthy food items, like confectionaries and processed snacks (21). However, it does not mandate selling healthy food items, nor does it provide guidance on the nutritional content of offered food and drink items (18). Additionally, although the MOH has made efforts to encourage reporting violations of school canteens, these initiatives do not appear to be exerting a positive impact on rising obesity rates amongst the youth of KSA. While policies and strategies exist, there is often insufficient follow-up and compliance monitoring to ensure they are effectively implemented.

Despite KSA’s efforts in legislating nutrition-related policies, such as implementing the largest global SSB tax at 50% (Alluhidan et al., 2022), the absence of evaluation or impact studies on school food environments or consumption patterns raises concerns about the effectiveness of these policies. Additionally, the Saudi Food and Drug Authority (SFDA) has introduced voluntary guidelines to restrict the marketing of unhealthy foods to children (29). However, evidence suggests that these guidelines have a limited impact. A recent content analysis found that unhealthy food items continue to be prominently advertised to children in KSA, particularly through social media platforms (30). This highlights the gap between policy and practice, due to voluntary compliance in social media platforms, where monitoring is challenging. Stronger enforcement of marketing restrictions is needed to reduce children’s exposure to unhealthy food marketing.

The MOE recommends incorporating PA into the school curriculum (24); however, this recommendation was not implemented in the schools assessed as part of this audit. Until 2018, physical education and PA were not formally part of the national curriculum for females, and cultural norms continue to restrict female participation. As noted by the principal of the high deprivation school, PA is often treated as a theoretical subject rather than a practical one. While PA is encouraged, it is not compulsory, and Saudi schools are likely falling short of global PA guidelines and recommendations, despite the increasing rates of obesity and physical inactivity among youth in KSA. Recent evidence revealed that 28.4% of Saudi adolescents are living with overweight or obesity (31), and 82.4**%** were classified as physically inactive (32). Another key finding from the school-based audit is the notable inequities in school resources, infrastructure, and health promotion practices across schools from different deprivation levels. While schools in medium and low deprivation areas had more developed infrastructure for PA, such as a dedicated space for sports in medium deprivation school, the facilities were often underutilized. The lack of adequate PA facilities in the high deprivation school, coupled with limited policies and resources for implementing health programs, create a challenge in ensuring equitable access to health resources across different deprivation levels. The absence of a formal health committee in the high deprivation school is an example of how lack of resources can hinder health promotion in general, and obesity prevention specifically within the school setting. Obesity prevention policies need to be supported by strong implementation to ensure effective improvement on student health outcomes (33). The school-based audit study also highlights the absence of a canteen in the high deprivation school. The financial circumstances of students led to the closure of the school shop in the high deprivation school, which contrasts with the functioning school shops in middle and low deprivation schools. However, even where food and drink shops existed, none of the schools provided healthy food options at affordable prices, which works as a barrier to promoting HE among students. Research indicates that the absence of affordable healthy food in schools can undermine efforts to combat childhood obesity, particularly in high deprivation schools where students may have limited resources and access to healthy food options (34), which was confirmed by the school-based audit.

The school-based audit revealed further inequities in canteen operations and pricing emerged. While some healthier options were present (milk in low deprivation school and date biscuits in middle deprivation school), affordability and consistency in pricing were major barriers. The variation in food pricing across schools highlights the disparities in accessing healthy food, with prices in the low deprivation school often closer to those found in local grocery stores, while those in middle deprivation schools were more expensive than grocery stores. In high-deprivation schools, the absence of a canteen restricted access to healthier options, which reflect health inequities. The lack of affordable, healthy options highlights the need for more standardized guidelines on what should be offered in school canteens.

Health promotion activities, such as healthy breakfast days, were also inconsistently implemented across the schools. The low deprivation school stood out for its proactive approach to health promotion, with various campaigns designed to raise students’ awareness of healthy behaviours. In contrast, the high deprivation school did not engage in such initiatives, pointing to gaps in the school’s approach to fostering a culture of health and well-being. These differences in health promotion practices across different socio-economic deprivation levels could contribute to disparities in health knowledge and behaviours among students, especially PA and HE behaviours.

This study highlights the importance of intersectoral collaboration in tackling childhood obesity. While initiatives like RASHAKA is highly commendable, no evaluation or impact studies exist. As noted by Al-Hazzaa and AlMarzooqi, many of KSA’s PA initiatives are fragmented, short-term, and inadequately evaluated (35), which aligns with the realities of policy implementation at the school level in KSA. To ensure effectiveness, obesity prevention strategies must be part of a comprehensive, multi-sectoral approach. Although KSA has legislated different obesity prevention policies and strategies, implementation at the school level remains inconsistent, inequitable, and fragmented. Without intersectoral collaboration, compliance monitoring, and attention to gender and socioeconomic inequities, national efforts to prevent adolescent obesity risk remaining aspirational rather than transformative.

As this study suggests, schools face significant challenges in implementing national obesity prevention policies within the school setting. With the absence of evaluation studies, it will be difficult to assess the impact or effectiveness of these policies. The national obesity-related strategies applicable to school settings emphasize the significance of changing the school food environment, incorporating health education, and offering opportunities for PA. The inclusion of school settings in many national programs in KSA demonstrates the possibility and importance of integrating obesity prevention initiatives into educational institutions.

Theoretical frameworks provide valuable lenses for interpreting these findings. The SEM emphasized how different levels of influences at the institutional and public policy levels interact to shape health behaviours. Without obesity prevention policy enforcement, however, interventions will likely fail to influence students’ behaviours effectively.

This study offers context-specific and novel insights into KSA’s school environments, which remains underexplored. To our knowledge, this is the first study to employ the culturally adapted ISCOLE questionnaire in KSA across schools of varying economic deprivation levels. The use of comprehensive search strategies to identify and verify obesity prevention policies (such as web-searching methods and hand-searching targeted websites) ensured a thorough search. Stakeholder consultations enhanced credibility through confirmation of findings. Combining a compliance study with an environmental school-based audit provided a perspective into policy intent and school-level implementation.

While this study offers valuable insights into the built environment of schools, it also has several limitations. First, it focuses on only three schools in Riyadh, which limits the generalizability of its findings to other regions or school types (for example: male-only schools, private schools, or primary schools). Although the ISCOLE was culturally adapted, further validation is required. Further research is needed to examine how schools in different regions of KSA are implementing obesity prevention strategies and contributing to national obesity prevention goals.

## 5. Conclusion

This study highlights gaps between national obesity prevention policies in KSA and their implementation in female high schools (practice). Challenges such as limited (or lack of) HE provision, inadequate PA facilities, and low canteen/shop adherence to healthy food checklists from the MOE (where available) restrict opportunities for adolescent girls to engage in healthy behaviours (HE and PA). This is compounded with inequities and disparities across schools from different levels of deprivation. Weak enforcement of policies and strategies, coupled with a lack of compliance monitoring, undermines the effectiveness of existing initiatives. This highlights the need for more intersectoral collaboration implementation of policies and improved monitoring and evaluation. Understanding the school context is critical for designing effective obesity prevention SBIs in this specific cultural context in the future. Without addressing these systemic gaps and challenges, national obesity prevention efforts are unlikely to achieve meaningful improvements to health behaviours or public health outcomes.

**Figure 1:**
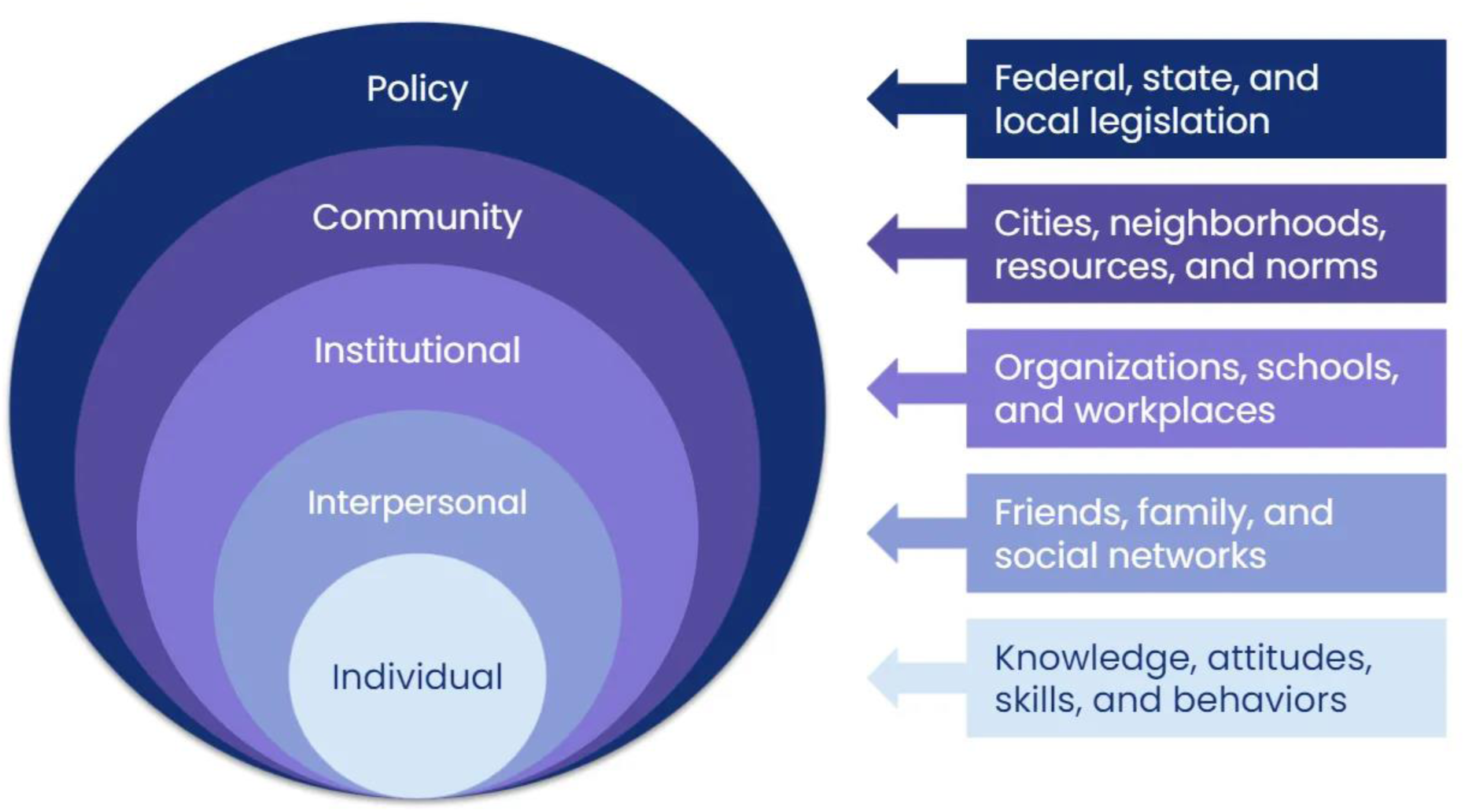
The Social Ecological Model (14)

## Data Availability

All data produced in the present study are available upon reasonable request to the authors.

## Acknowledgements

We thank the participating schools and staff for their valuable contributions. We thank Dr Hannah O’Hara for checking the final manuscript.

## Authorship

SA, LM and JW designed the study; SA conducted the study, collected and analysed data, and drafted the initial manuscript. LM and JW contributed to interpretation. JW and KA supervised study design and JW and LM provided critical revisions. All authors read and approved the final manuscript.

## Ethical Standards Disclosure

This study was conducted according to the guidelines laid down in the Declaration of Helsinki and all procedures involving research study participants were approved by Princess Nourah bint Abdulrahman University’s institutional review board, Saudi Ministry of Education, and Queen’s University Belfast’s research ethics committee (MHLS 24_47). Written informed consent was obtained from all subjects.

