## Supplementary materials 1 for "Using the Social Ecological Model to explore policy and institutional aspects of the school environment in relation to healthy eating and physical activity: An audit of female-only high schools in the Kingdom of Saudi Arabia"

**Supplementary File S1: ISCOLE school environment questionnaire**

### ISCOLE SCHOOL ENVIRONMENT QUESTIONNAIRE

#### A. SCHOOL CHARACTERISTICS

**1. What is your position at this school?** Principal Vice Principal Teacher Other:

| | | |
|---|---|---|
| 2. | **What is the total number of students in your school?** (Please estimate) | ____ students |
| 3. | **What is the total number of teachers (full time equivalents) in your school?** (Please estimate) | _ teachers |
| 4. | **What grades are taught at your school?** | _ to |
| 5. | **How many days (excluding holidays) do your students attend school during the academic school year?** | |
| B. | **POLICIES AND PRACTICES** | |

For the following section, "policies" refers to any mandates issued by the state, the local school board, or any other agency, including policies developed by your school or (district/diocese), that affects your school environment and that have been officially adopted by your school or district. This section also asks about practices (what your students and staff are allowed to do on a regular basis) that you might follow to promote the health and well-being of students.

1. **Does your school have written policies or practices concerning physical activity?**

- ☐ **Yes**, existing written policies
- ☐ **Yes**, written policies still under development
- ☐ **Yes,** practices
- ☐ No
- ☐ N/A

***If yes, what are the policies/practices?***

2. **Does your school have written policies or practices concerning <u>healthy eating</u>?**

- ☐ **Yes**, existing written policies
- ☐ **Yes**, written policies still under development
- ☐ **Yes,** practices
- ☐ No
- ☐ N/A

***If yes, what are the policies/practices?***

3. **Does your school have a committee that oversees or offers guidance on the development of policies and practices concerning physical activity and healthy**

**eating at your school (e.g., health action team, school health or wellness council)?**

- **Yes**, both physical activity and healthy eating
- **Yes**, physical activity only
- **Yes,** healthy eating only
- No

**C. PHYSICAL ACTIVITY**

4. **What percent of students participate in the following extracurricular activities offered by your school?**

(Please estimate)

*Not available*

*Less than 10%*

*10-24% 25-49% 50%+*

***Does the sports (PA) offered to students differ across different grades?***

5. **Does your school offer late bus/transportation service to students who participate in extra-curricular activities?**

Yes No

6. **From the following list, please indicate which sports (PA) are offered in your school available to students in grades 10-11-12:**

 a. Not applicable, school does not offer interschool athletics to students.

| | | | | | |
|---|---|---|---|---|---|
| b. Basketball. | c. Volleyball | d. Soccer | e. Football | f. Baseball | g. Gymnastics |
| h. Wrestling | i. Badminton | j. Swimming | k. Other | | |

**For the following questions, please consider students in grades 10-11-12 when answering.**

7. **How many breaks of 15 to 29 minutes do students have in a day?**

zero 1 2 3 or more

8. **How many breaks of 30 minutes or more do students have in a day?**

zero 1 2 3 or more

9. **How much class time is mandated by the Ministry of Education to be allotted to physical education (PE)/Daily Physical Activity (DPA) for students?**

**minutes per** *[check the box indicating the time unit]* **week ** **day **

 No specific amount is mandated

..............

10. **Compared to the class time allotted to physical education (PE)/Daily Physical Activity for students 4 as mandated by the ministry of Education, do students in your school receive on average:**

 Less than the mandated amount

 Approximately the mandated amount

 More than the mandated amount

 No specific amount is mandated

11. **To the best of your knowledge, how well do each of the following statements characterize your school?**

*A lot* *Some* *Very little*

*Not at all*

*Don't know*

a. We use physical activity as a reward

b. We promote physical activity during or as part of special events

| |
|---|
| c.We integrate physical activity into other curriculum areas |
| d. We use physical activity as a punishment for bad behavior (e.g., withholding recess, administering push-ups or laps). |

12. **What kind of transportation do students use to arrive to their school?**

a. Car
b. Bus
c. Other

**D. SCHOOL FACILITIES**

13. **Do the majority of students at your school have regular access to any of the following during school hours*?**

*During school hours means from the first bell to the last bell, including both instructional and non-instructional time (e.g., lunch). *(**If YES**, please use the checklist below. **If NO**, skip to question 19)*

a. Gymnasium

b. Other large room suitable for physical activity (e.g., auditorium, cafeteria)

*Yes, on*

*grounds Only*

*Yes, off grounds only*

*Yes, both on*

*and off* *No*

*grounds*

*Don’t know*

d. Running track

f. Outdoor paved area suitable for sports (PA)

h.Indoor swimming pool

| |
|---|
| i.Secure change room lockers available for use during physical activity |
| j.Change rooms available for use before and after physical activity |
| k.Showers available for use before or after physical activity |
| l.Bicycle racks |
| m. **<u>If yes</u>**, are the racks in a secure area to avoid theft? |
| n.Grassy playground area |
| o.Playground equipment (e.g., climbing structures, swings) |
| p.Art room |
| q. Music room |

14. **Do students have access to the following facilities where they can buy foods or drinks?**

*Yes* *No*

f. Milk vending machine/ milk program (e.g., milk, chocolate milk)

15. ***Outside of school hours*,*** **does your school permit regular student access to the following?**

**Outside of school hours means before and/or after school, evenings and weekends. Student access may occur via school-led, community-led or informal use.*

| | *Yes* | *No* | *Don't know* | *N/A* |
|---|---|---|---|---|
| a. Gymnasium | | | | |
| b. Indoor facilities | | | | |
| c.Outdoor facilities (e.g., playing fields, paved activity areas, baseball diamond) | | | | |
| d. Equipment (e.g., basketballs) | | | | |

16. ***Outside of school hours****, **does your school allow community groups to use the school facilities?**

**Outside of school hours means before and/or after school, evenings and weekends.*

Yes  No  Don't know 

**E. HEALTHY EATING**

17. **Does your school provide any of the following to promote the sale of healthy food**? (Check all that apply)

| | *Cafeteria* | *Snack bar/ School shop* |
|---|---|---|
| *Vending machine(s)* | | |
| a. Healthy food choices at a reasonable/subsidized price | | |
| b. Healthy eating promotional materials (e.g., posters) | | |
| c.Daily healthy eating specials | | |
| d. Healthy eating cafeteria program (e.g., Eat Smart or independent program) | | |

18. **Does your school ensure that all students, regardless of ability to pay, have access to fruits and vegetables?**

- Yes, entire school year
- Yes, occasional/short term
- No

19. **Does your school offer any of the following? (Check all that apply)**

- Cooking classes
- Gardening (e.g., growing produce)
- Field trips to farms/farmers' markets
- Media literacy on special topics related to healthy eating (e.g., body image, eating disorders)
- Field trips to the local grocery store

20. **During the past 12 months, did your school initiate/continue any of the following activities/programs at your school?**

| | *Yes* | *No* | *N/ A* |
|---|---|---|---|
| a. Offered healthy food choices during breakfast program | | | |
| b. Offered healthy food choices during lunch program | | | |
| c.Offered healthy food choices in the cafeteria(s) | | | |
| d. Offered healthy food choices in the snack bar/school shop(s) | | | |
| e. Offered healthy food choices in the vending machine(s) | | | |
| f.Organised Nutrition Month activities | | | |
| g. Stopped the sale of junk food | | | |
| h. Held junk food free days | | | |
| i.Stopped the sale of sugar-sweetened beverages | | | |

**F. NEIGHBORHOOD/COMMUNITY**

21. **How much of a problem are the following in the neighborhood where this school is located?**

*Major*

*problem*

*Moderate problem*

*Minor problem*

*Not a problem*

*I don't know*

| |
|---|
| a.Tensions based on racial, ethnic, or religious differences |
| b.Garbage, litter, or broken glass in the street or road, on the sidewalks, or in yards |
| d. Selling or using drugs in public |
| d.Gangs |
| e.Heavy traffic |
| f.Vacant or shabby houses and buildings |
| g.Crime in the neighborhood |

Supplementary Table S2: The MOE Health Regulations for School Canteens checklist

- *Unallowed food items in Saudi school canteens*

| Food Item | Present/Not Present | Observational Notes |
|---|---|---|
| Soft drinks/soda | | |
| Energy drinks | | |
| Iced tea | | |
| Flavored water | | |
| Juices with less than 30% natural juice/nectar juices | | |
| Flavored yogurt/milk | | |
| Meat (fish, chicken, beef) | | |
| Liver | | |
| Hot dog/sausage/mortadella | | |
| Falafel | | |
| Custard/chocolate/vanilla pastries | | |
| Fried food | | |
| Potato chips | | |
| Salted nuts | | |
| Ice cream | | |
| Popcorn | | |
| Candy | | |
| Bubble gum | | |
| Cheese/corn puffs | | |
| Chocolates in any form | | |
| Pickles | | |
| Mayonnaise | | |

| |
|---|
| Donuts |
| Croissants |
| Food items containing monosodium glutamate |

- *Recommended food items in Saudi school canteens*

| Food items | Present/Not Present | Observational notes |
|---|---|---|
| Full fat/low fat fresh milk | | |
| Full fat/low fat long-life milk | | |
| Unflavored natural yogurt/yogurt with fresh fruits | | |
| Bottled water | | |
| 100% natural juice | | |
| Juices with more than 30% natural juice | | |
| Gluten-free sandwiches for celiac students | | |
| Sandwiches restricted to the following fillings: (cheese/labneh/zaatar/honey/jam/egg /peanut butter/egg/fava beans/hummus) | | |
| Fruits (fresh/dried) | | |
| Fresh vegetables | | |
| Packaged dates | | |
| Packaged date/fig biscuits (mamoul) | | |
| Gluten-free biscuits | | |
| Unsalted nuts | | |

Supplementary Figure S3: Ministry of Health healthy breakfast infographic

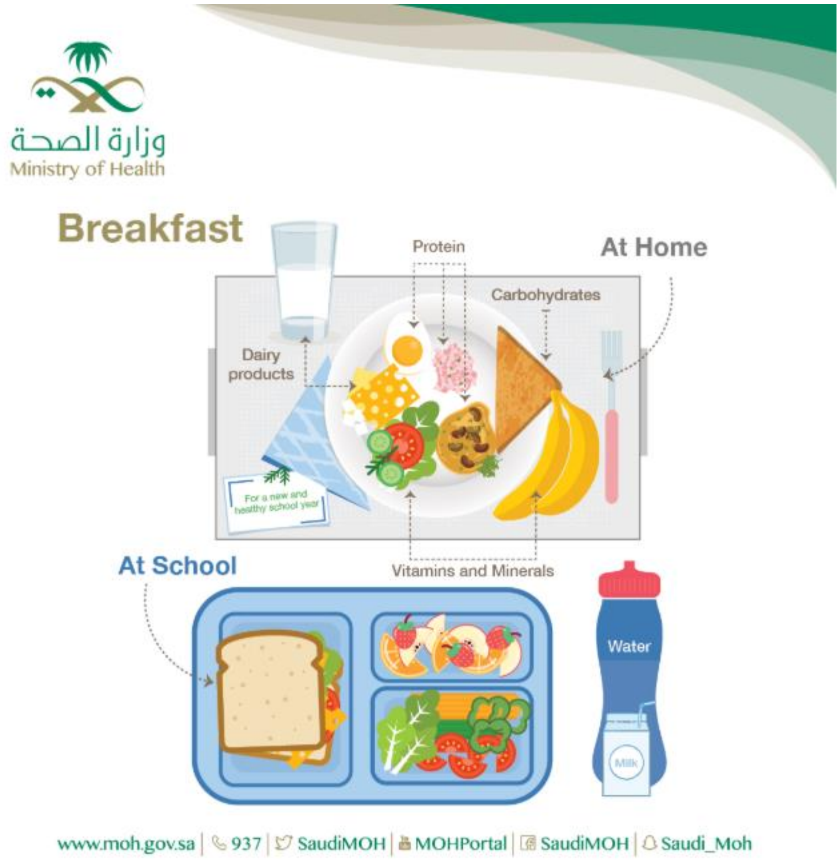

Supplementary Figure S4: Photographs of low deprivation school canteen

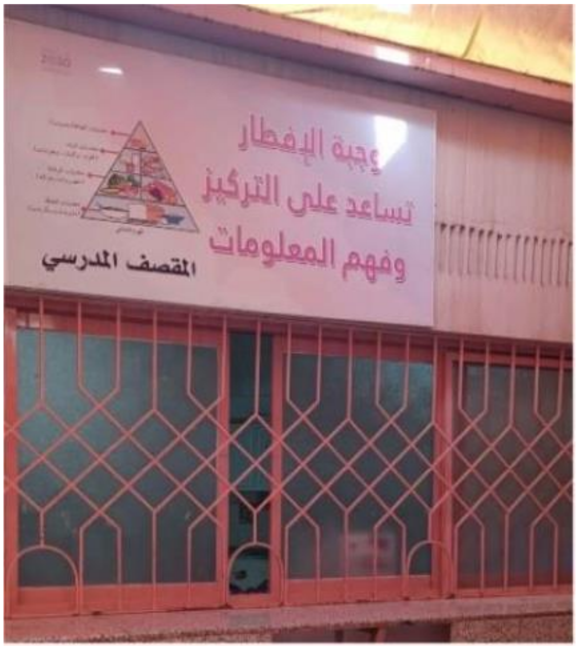

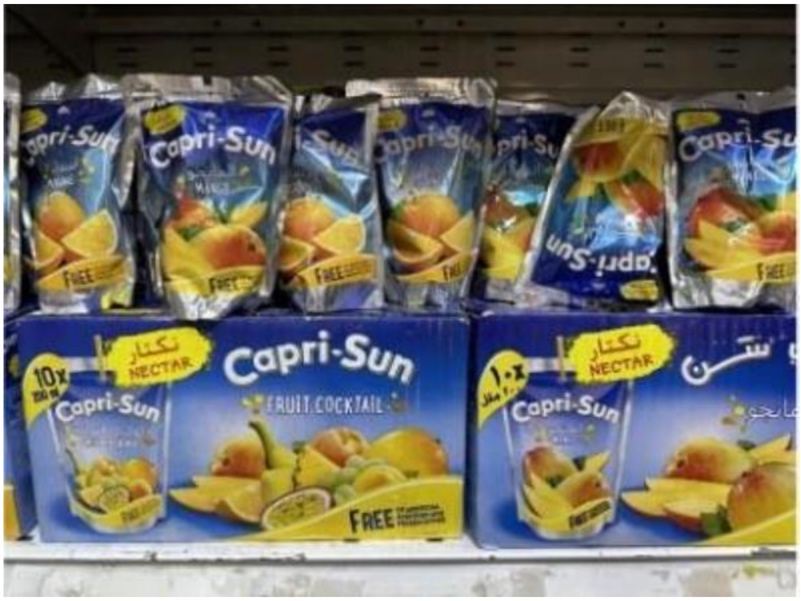

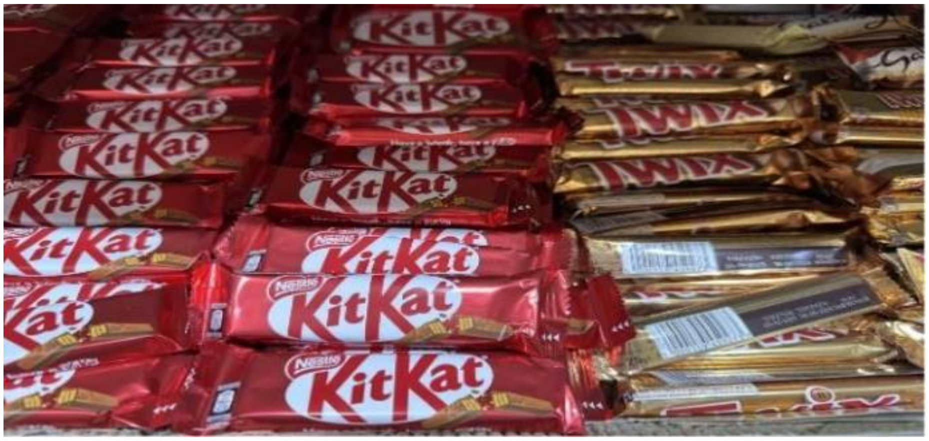
KitKat
Nestlé
TWIX
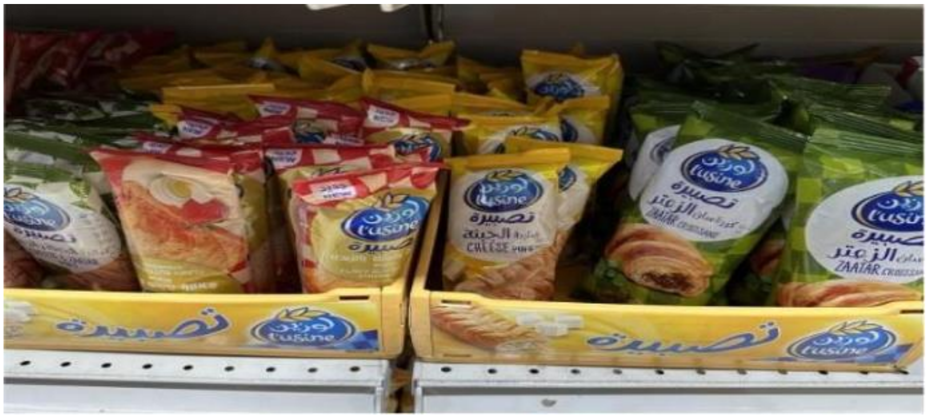
L'usine
تصبيرة
الزعتر
ZAATAR
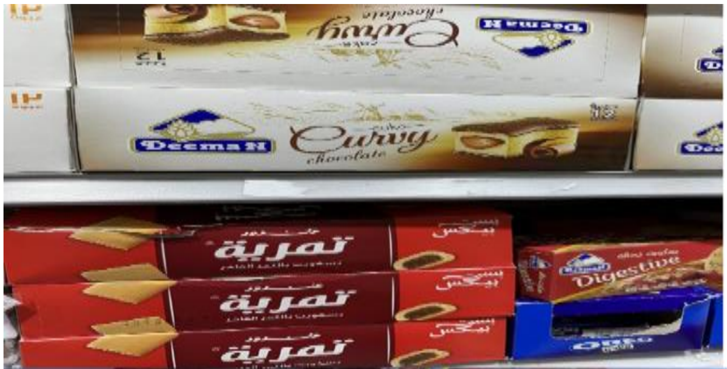
Deemah
Curvy
chocolate
تمرية
Digestive
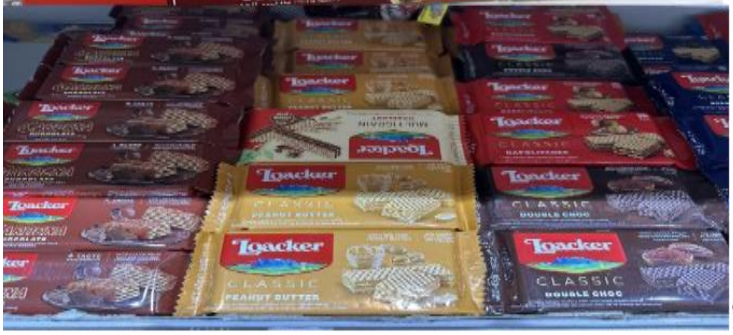
Loacker
CLASSIC
PEANUT BUTTER
DOUBLE CHOC

Supplementary Figure S5: Photographs of middle deprivation school PA facilities

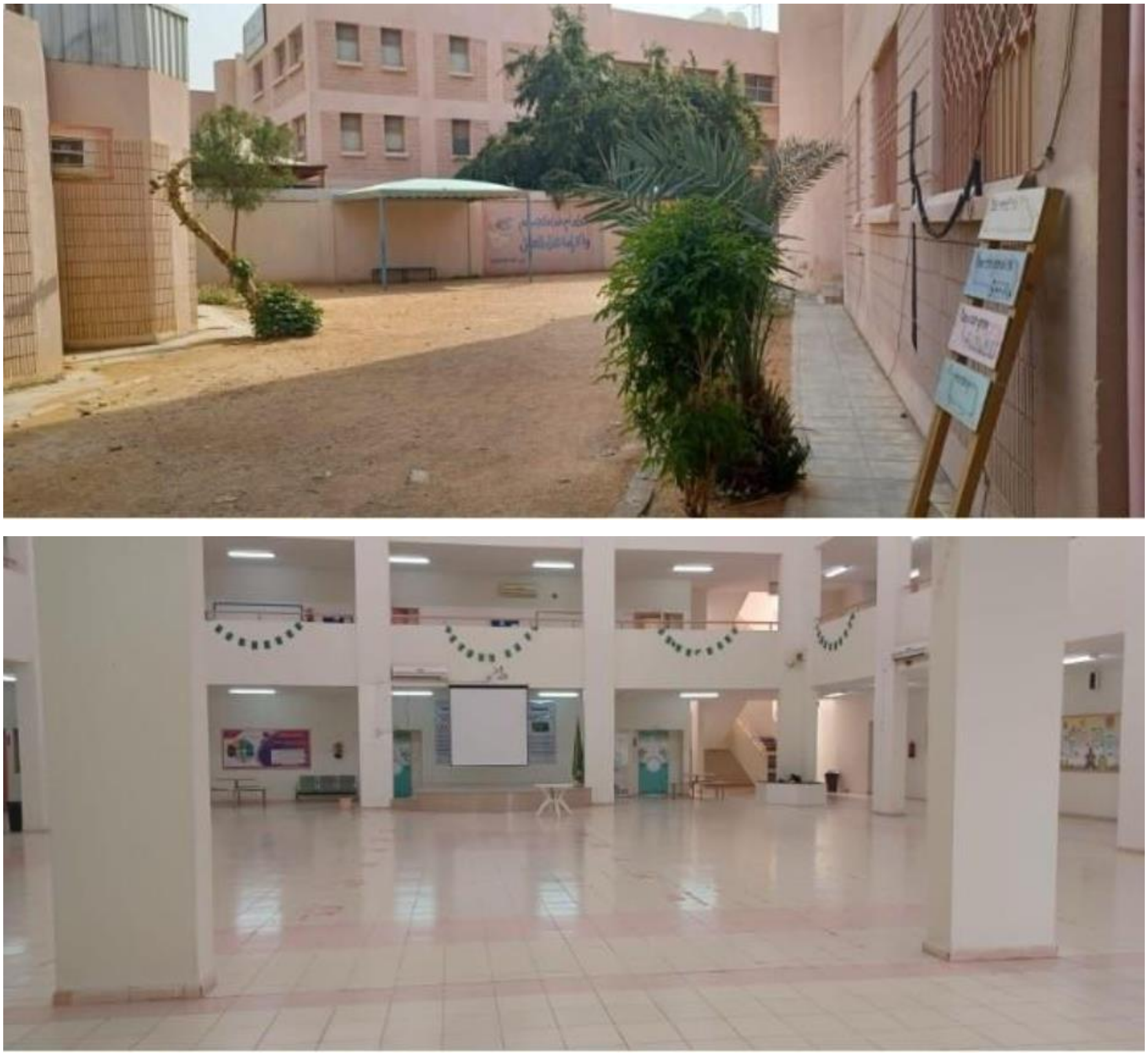

Supplementary Figure S6: Photographs of middle deprivation school canteen

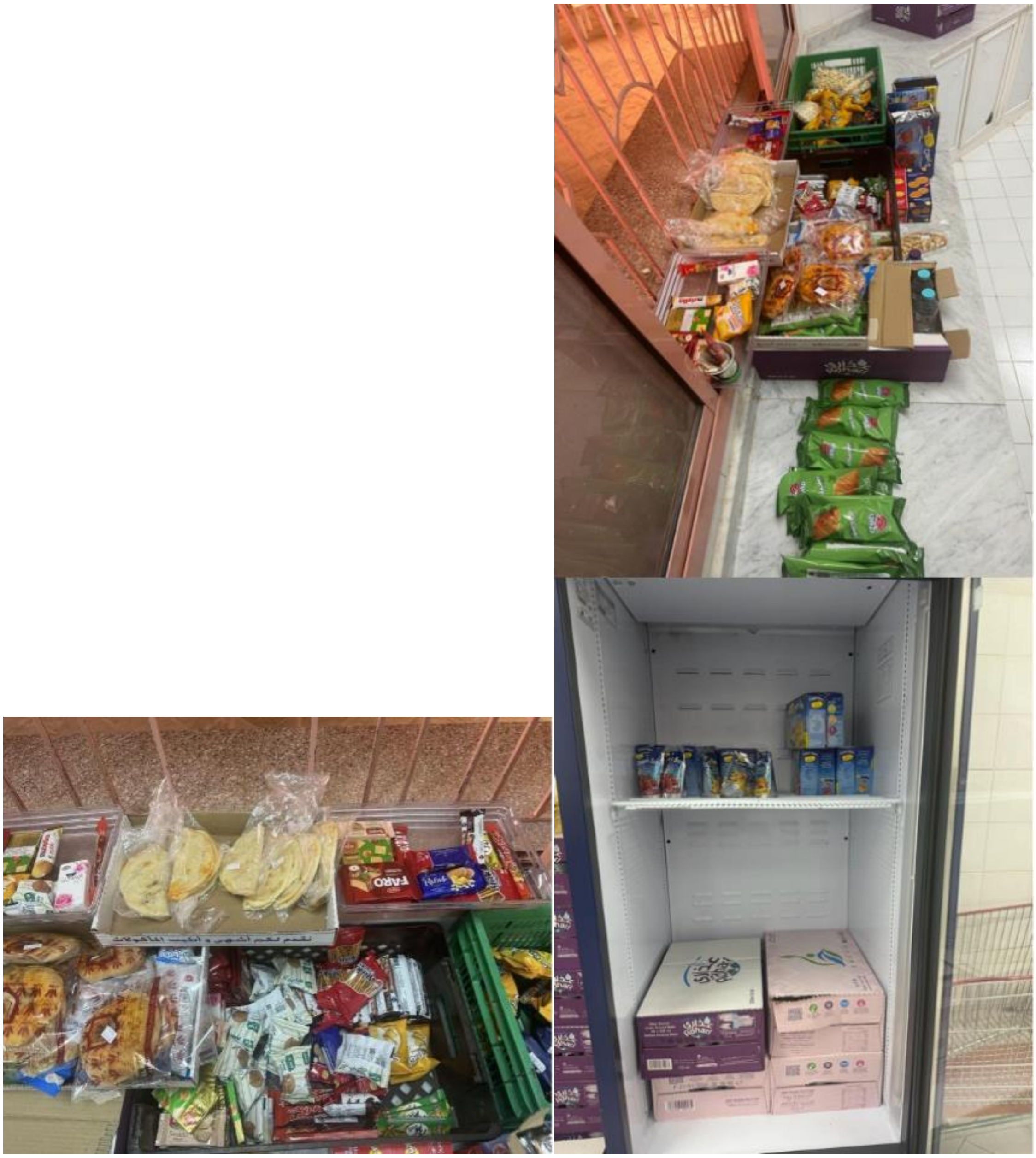

Supplementary Figure S7: Photographs of Middle deprivation school PA facilities

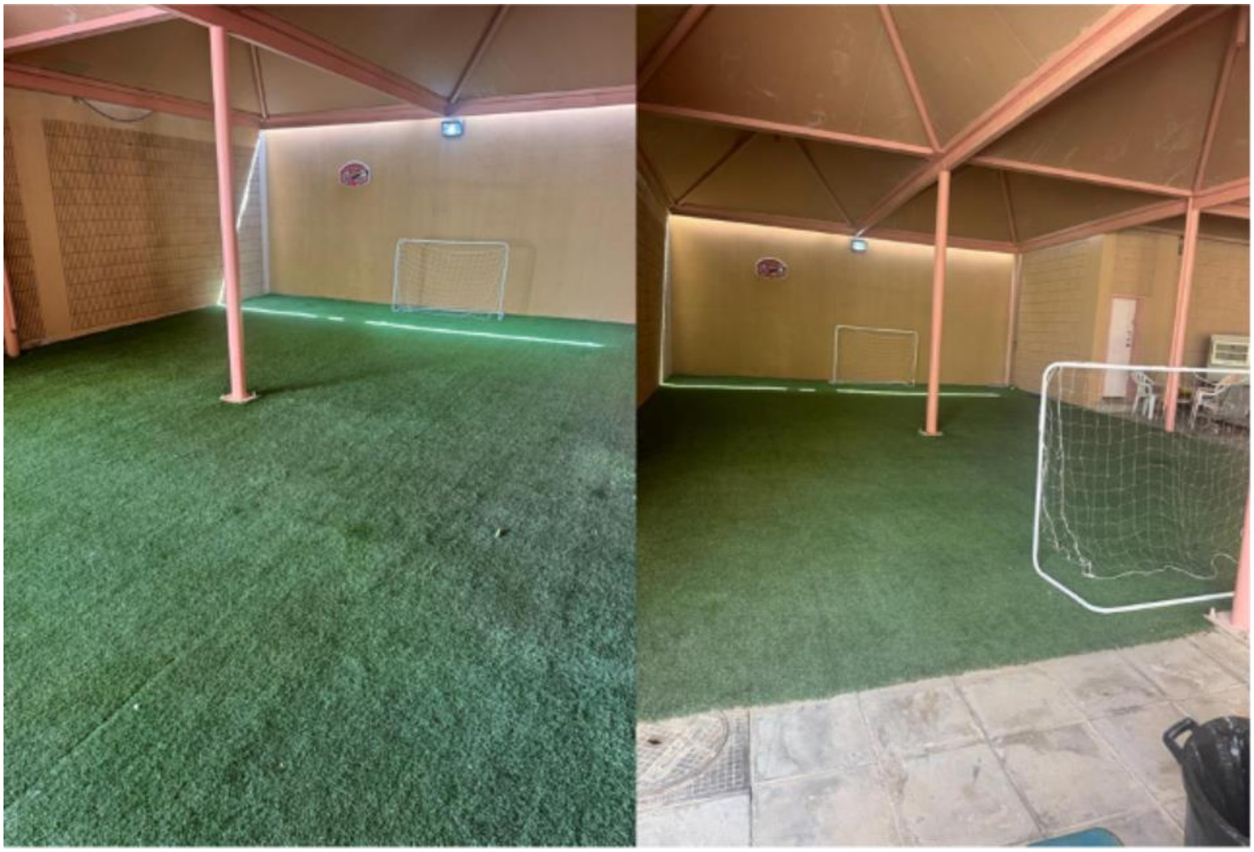
